# Comparison of primer-probe sets among different master mixes for laboratory screening of Severe Acute Respiratory Syndrome - Coronavirus 2 (*SARS-CoV-2*)

**DOI:** 10.1101/2020.06.20.20136242

**Authors:** Hoang Quoc Cuong, Nguyen Duc Hai, Hoang Thuy Linh, Nguyen Hoang Anh, Nguyen Trung Hieu, Cao Minh Thang, Nguyen Thi Thanh Thao, Phan Trong Lan

## Abstract

**Background:** There is a shortage of chemical reagents for severe acute respiratory syndrome - coronavirus 2 (*SARS-CoV-2*) diagnosis and a surge of *SARS-CoV-2*cases, especially in limited-resource settings. Therefore, the combination of an optimal assay kit is necessary.

**Methods:** We compared the ability to screen *SARS-CoV-2* among three primer-probe sets in two different master mixes, Invitrogen™ SuperScript™ III One-Step RT-PCR and LightCycler Multiplex RNA Virus Master.

**Results:** The assay with TIB-Molbiol, IDT, and Phu Sa sets for LightCycler Multiplex RNA Virus Master or Invitrogen™ SuperScript™ III One-Step RT-PCR showed positive results from a single reaction of triplicate in the three days of 4.8 copies per reaction. R-squared and amplification efficiency were 0.97 and ranged from 107 to 108%, respectively.

**Conclusions:** Our findings indicated that TIB-Molbiol, IDT, and Phu Sa primer-probe sets could be beneficial for the laboratory screening of *SARS-CoV-2* by RT-qPCR assay of E gene. There is a need to consider the combination of these reagent sets as a new strategy to increase the testing capacity of screening programs for COVID-19.

## 1. Introduction

Severe Acute Respiratory Syndrome Coronavirus 2 (*SARS-CoV-2*) has a threat to human health which involves over 7,273,958 confirmed cases and 413,372 deaths [1]. While waiting for the coronavirus vaccine approval, molecular testing for *SARS-CoV-2* is one of the important strategies to prevent and reduce the rate of infection by case identification, isolation, social distancing, and proper treatment [2, 3].

Although many factors are leading to the low sensitivity of *SARS-CoV-2* such as *a)* detection depends on the location of clinical specimens, *b)* low patient viral load, *c)* sporadic shedding, and *d)* discrepancy in detection kits from various producers [4, 5]. However, the molecular diagnosis of *SARS-CoV-2* using RT-qPCR assay is a gold standard method [6-8]. Consequently, the combination of an optimal assay kit is necessary because of the shortage of chemical reagents for *SARS-CoV-2* diagnosis and the surge of *SARS-CoV-2* cases, especially in limited-resource settings.

In the present study, we aim to analyze the commonly used primer-probe sets, targeting E gene of *SARS-CoV-2* by the RT-qPCR assay for laboratory screening to increase testing capacity in the context of thousands of overseas travelers returning to their countries.

## 2. MATERIALS AND METHODS

### 2.1. Primer-probe information

In this study, these three primer-probe sets based on the sequence information received from three different companies, TIB-Molbiol (Berlin, Germany), IDT (Integrated DNA Technologies, Skokie, Illinois, USA) and Phu Sa (Phu Sa Biochem, Vietnam) were used for comparative analysis [8] *(Supplementary Table* 1).

### 2.2. Viral preparation

The infection assays were performed in a biosafety level 3 laboratory. Vero E6 cell lines were infected with a clinical isolate *SARS-CoV-2* [9]. After 72 hours, the virus medium was inactivated at 65°C for 1 hour. Viral RNA was then isolated from the culture medium using the QIAamp viral RNA extraction Kit (Qiagen, Hilden, German) following the manufacturer’s instructions. The copy number of RNA extracted from *SARS-CoV-2* strain was estimated through a standard curve, which was published in a previous study [10].

*SARS-CoV-2* strain isolated in this study is MT192773. The length of the genome sequence was 29,890 bp without gaps and high coverage 1,897×. This strain belonged to Betacoronavirus B type and of 99.98% sequence similarity at the nucleotide level which was isolated in Wuhan (MT019529), and >90.56% similarity with *SARS-CoV* isolated from pangolin (EPIISL410721). There have been four mutations classified as nonsynonymous mutations such as G8388A (serine to asparagine), A8987T (isoleucine to phenylalanine), and C10232T (arginine to cysteine) and a synonymous mutation is G3778A, which was published in a previous study [9].

### 2.3. Real-time reverse transcription-polymerase chain reaction (RT-PCR) assay confirmation for *SARS-CoV-2*

RNA extracted specimens from the inactivated virus were tested for comparative assay of *SARS-CoV-2* by RT-qPCR on a LightCycler 480 or ABI 7500 system following the manufacturer’s protocol (Invitrogen™ SuperScript™ III One-Step RT-PCR System or LightCycler Multiplex RNA Virus Master). In this study, the reaction combination was prepared by multiplying the volumes of each reagent in Table 1.

**Table 1.**
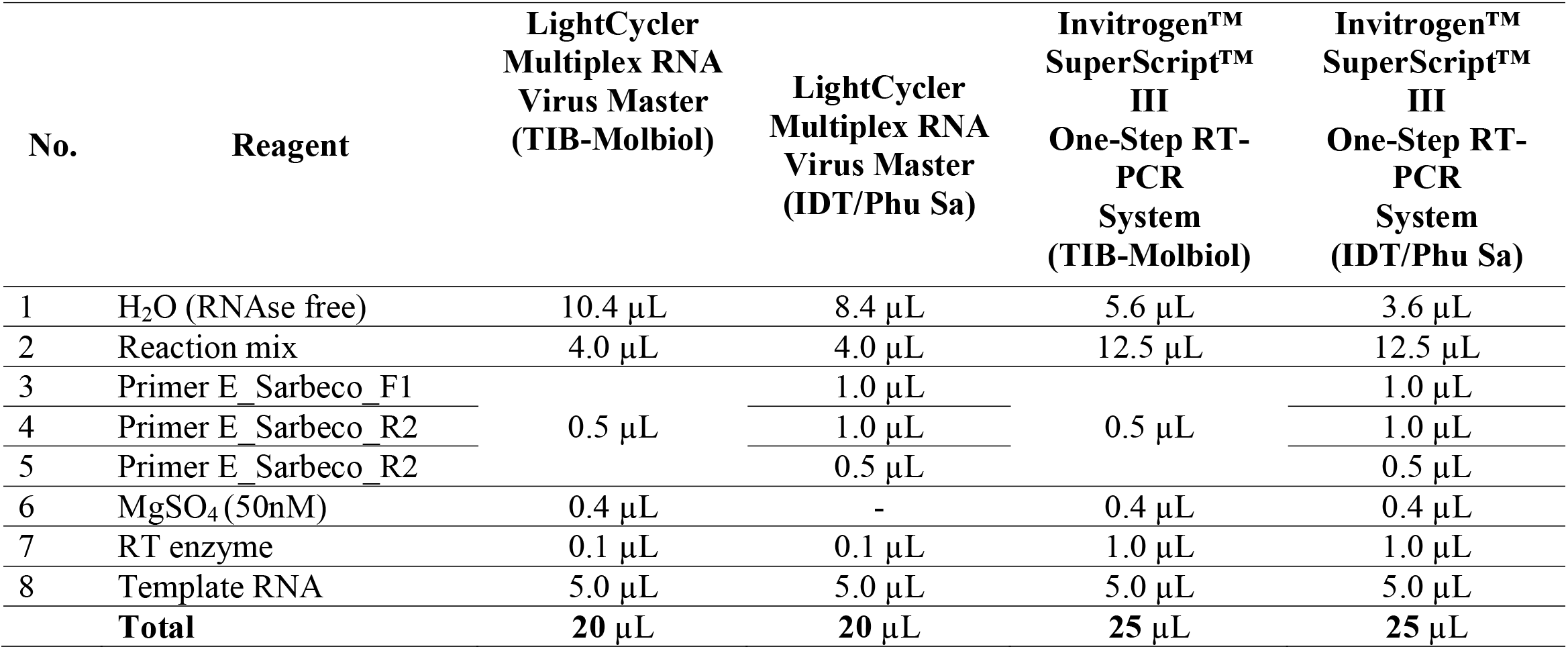
Volumes of reagents for reactions using two different polymerase enzymes combined with three primer-probe sets.

RT-qPCR conditions applied in the present study with details described in Table 2. A cycle threshold value (Ct-value) of ≥40 was defined as a negative test [8].

**Table 2.**
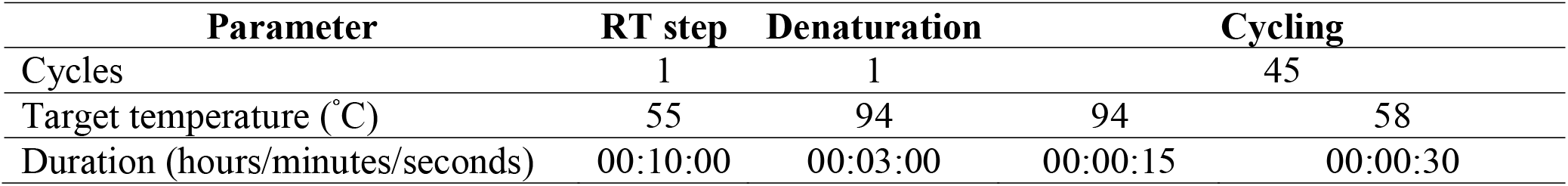
RT-qPCR conditions applied in the present study.

| Parameter | RT step | Denaturation | Cycling |  |
| --- | --- | --- | --- | --- |
| Cycles | 1 | 1 | 45 |  |
| Target temperature (°C) | 55 | 94 | 94 | 58 |
| Duration (hours/minutes/seconds) | 00:10:00 | 00:03:00 | 00:00:15 | 00:00:30 |

### 2.4. Analysis

In this study, Data were entered using Epi-Data version 3.1 (EpiData Association, Odense, Denmark, 2005), and all statistical analyses were performed using Stata version 13.0 (StataCorp, TX, 2013).

The results were summarized using means and standard deviation (SD) for continuous variables. Linear regression analysis was performed to estimate the R-square. Amplification efficiency (AE) was calculated using the equation AE = -1+10^(−1/slope)^ [11].

### 2.5. Ethical statement

The study protocol has been reviewed and ratified by the Pasteur Institute Ho Chi Minh City Institutional Review Board (reference number: 433/XN-PAS).

## 3. RESULTS

In this study, the assay with TIB-Molbiol, IDT, and Phu Sa sets for LightCycler Multiplex RNA Virus Master showed positive results from a single reaction of triplicate in the three days of 4.8 copies/reaction (Table 3).

**Table 3.**
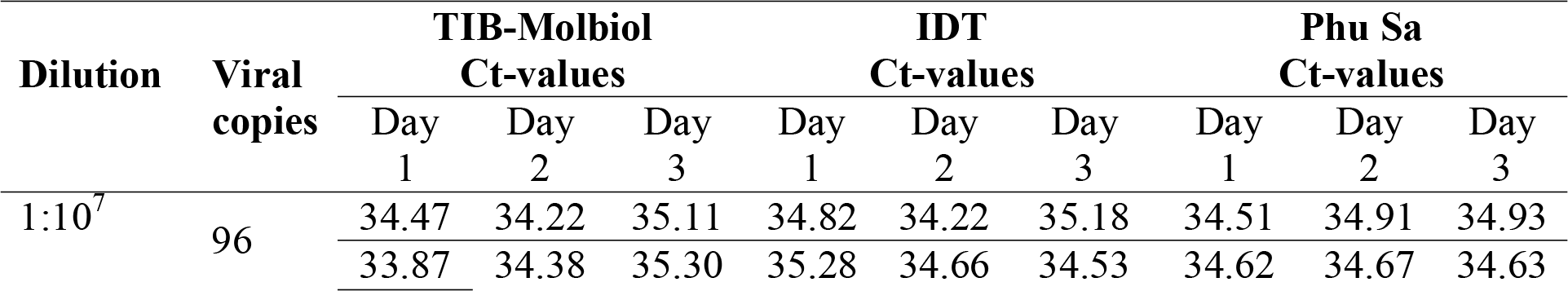
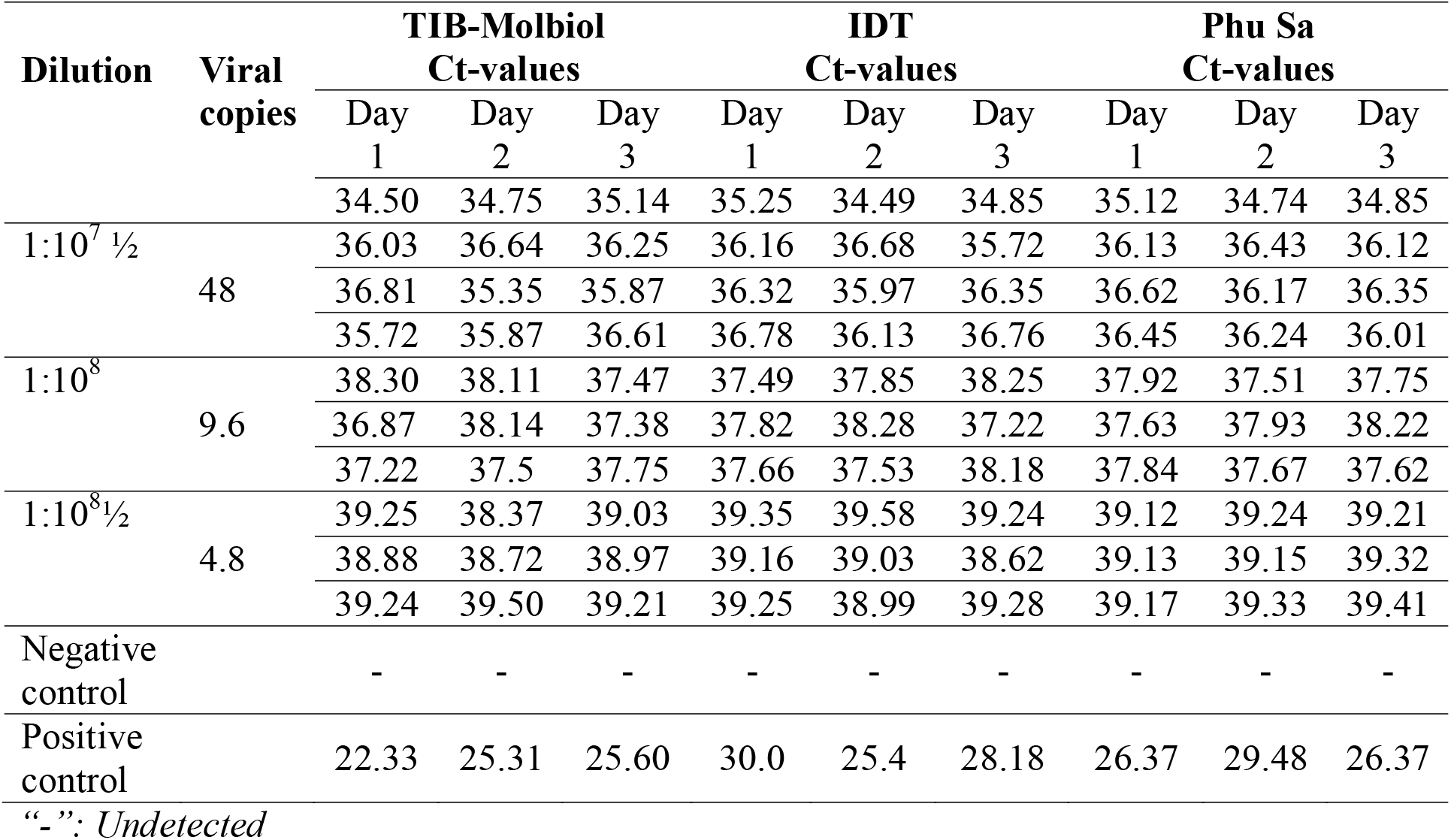
The results of LightCycler Multiplex RNA Virus Master with three primer-probe sets (IDT, Phu Sa, TIB-Molbiol).

The Ct-values (mean±SD) of E gene (TIB-Mobiol), IDT, and Phu Sa at 1:10^8^½ were 39.02±0.34, 39.17±0.34 and 39.23±0.10, respectively. R^2^ value from TIB-Mobiol, IDT, and Phu Sa showed equal values of 0.97. Similarly, the AE of each set was also showing the same value, for the figures of TIB-Mobiol and IDT were 107; and Phu Sa was 108 (Table 4).

**Table 4.** Comparison of Ct-value means for LightCycler Multiplex RNA Virus Master with three primer-probe sets (IDT, Phu Sa, TIB-Molbiol).

| Dilution | Viral<br>copies | TIB-Molbiol |  |  |  | IDT |  |  |  | Phu Sa |  |  |  |
| --- | --- | --- | --- | --- | --- | --- | --- | --- | --- | --- | --- | --- | --- |
|  |  | Ct-values |  |  |  | Ct-values |  |  |  | Ct-values |  |  |  |
|  |  | Mean | SD | R<sup>2</sup> | AE | Mean | SD | R<sup>2</sup> | AE | Mean | SD | R<sup>2</sup> | AE |
| 1:10<sup>7</sup> | 96 | 34.64 | 0.47 | 0.97 | 107 | 34.81 | 0.37 | 0.97 | 107 | 34.78 | 0.19 | 0.97 | 108 |
| 1:10<sup>7 1/2</sup> | 48 | 36.13 | 0.49 |  |  | 36.32 | 0.36 |  |  | 36.28 | 0.20 |  |  |
| 1:10<sup>8</sup> | 9.6 | 37.64 | 0.47 |  |  | 37.81 | 0.37 |  |  | 37.79 | 0.22 |  |  |
| 1:10<sup>8 1/2</sup> | 4.8 | 39.02 | 0.34 |  |  | 39.17 | 0.27 |  |  | 39.23 | 0.10 |  |  |

The assay with TIB-Molbiol, IDT and Phu Sa primer-probe sets for the Invitrogen™ SuperScript™ III One-Step RT-PCR System exhibited positive results from a single reaction of triplicate in the three days of 4.8 copies/reaction (Table 5). Furthermore, these three primer-probe sets showed the equivalent sensitivity in low concentrations for LightCycler Multiplex RNA Virus Master.

**Table 5.** The results of Invitrogen™ SuperScript™ III One-Step RT-PCR System with three primer-probe sets (IDT, Phu Sa, TIB-Molbiol).

| Dilution | Copies/<br>reaction | TIB-Molbiol<br>Ct-values |  |  | IDT<br>Ct-values |  |  | Phu Sa<br>Ct-values |  |  |
| --- | --- | --- | --- | --- | --- | --- | --- | --- | --- | --- |
|  |  | Day<br>1 | Day<br>2 | Day<br>3 | Day<br>1 | Day<br>2 | Day<br>3 | Day<br>1 | Day<br>2 | Day<br>3 |
| 1:10<sup>8</sup> | 96 | 34.23 | 34.51 | 34.81 | 35.07 | 34.91 | 35.03 | 34.34 | 35.21 | 35.31 |
|  |  | 34.16 | 34.66 | 34.6 | 35.15 | 33.94 | 34.34 | 35.03 | 34.87 | 34.62 |
|  |  | 34.09 | 34.09 | 34.83 | 35.12 | 34.82 | 34.21 | 34.24 | 34.86 | 35.32 |
| 1:10<sup>8</sup> ½ | 48 | 36.31 | 35.72 | 36.16 | 36.53 | 35.44 | 35.84 | 35.84 | 36.36 | 36.12 |
|  |  | 35.58 | 35.58 | 36.34 | 36.64 | 36.41 | 36.34 | 36.81 | 36.53 | 35.74 |
|  |  | 35.69 | 36.00 | 36.14 | 36.58 | 35.71 | 36.66 | 36.71 | 36.34 | 36.83 |
| 1:10<sup>9</sup> | 9.6 | 37.09 | 37.83 | 37.16 | 36.94 | 37.34 | 37.91 | 37.34 | 38.32 | 38.31 |
|  |  | 37.23 | 37.09 | 37.71 | 38.03 | 38.07 | 38.15 | 37.86 | 38.21 | 37.87 |
|  |  | 37.51 | 37.66 | 37.6 | 38.1 | 37.82 | 37.19 | 38.03 | 37.62 | 37.24 |
| 1:10<sup>9</sup> ½ | 4.8 | 38.59 | 38.59 | 38.66 | 39.62 | 39.32 | 38.82 | 38.82 | 39.72 | 38.74 |
|  |  | 39.33 | 38.63 | 39.1 | 38.42 | 38.71 | 39.51 | 39.77 | 39.59 | 39.52 |
|  |  | 39.16 | 39.01 | 39.31 | 39.46 | 39.41 | 39.55 | 39.36 | 39.12 | 39.37 |
| Negative control |  | - | - | - | - | - | - | - | - | - |
| Positive control |  | 24.27 | 25.91 | 22.81 | 28.40 | 28.38 | 26.72 | 20.81 | 30.56 | 29.39 |

“-”: Undetected

In this study, we found that the R-square values from TIB-Molbiol, IDT and Phu Sa were the same (0.97). Similar to the results of LightCycler Multiplex RNA Virus Master, the AE of IDT, and Phu Sa was also the same value (108); and 107 for TIB-Molbiol (Table 6).

**Table 6.** Comparison of Ct-value means for Invitrogen™ SuperScript™ III One-Step RT-PCR System with three primer-probe sets (IDT, Phu Sa, TIB-Molbiol).

| Dilution | Viral<br>copies | TIB-Molbiol<br>Ct-values |  |  |  | IDT<br>Ct-values |  |  |  | Phu Sa<br>Ct-values |  |  |  |
| --- | --- | --- | --- | --- | --- | --- | --- | --- | --- | --- | --- | --- | --- |
|  |  | Mean | SD | R<sup>2</sup> | AE | Mean | SD | R<sup>2</sup> | AE | Mean | SD | R<sup>2</sup> | AE |
| 1:10<sup>7</sup> | 96 | 34.44 | 0.30 | 0.97 | 107 | 34.73 | 0.45 | 0.97 | 108 | 34.87 | 0.39 | 0.97 | 108 |

| Dilution | Viral<br>copies | TIB-Molbiol |  |  |  | IDT |  |  |  | Phu Sa |  |  |  |
| --- | --- | --- | --- | --- | --- | --- | --- | --- | --- | --- | --- | --- | --- |
|  |  | Ct-values |  |  |  | Ct-values |  |  |  | Ct-values |  |  |  |
|  |  | Mean | SD | R<sup>2</sup> | AE | Mean | SD | R<sup>2</sup> | AE | Mean | SD | R<sup>2</sup> | AE |
| 1:10<sup>7</sup> ½ | 48 | 35.95 | 0.31 |  |  | 36.24 | 0.44 |  |  | 36.36 | 0.40 |  |  |
| 1:10<sup>8</sup> | 9.6 | 37.43 | 0.29 |  |  | 37.73 | 0.45 |  |  | 37.87 | 0.40 |  |  |
| 1:10<sup>8</sup> ½ | 4.8 | 38.93 | 0.30 |  |  | 39.20 | 0.43 |  |  | 39.33 | 0.37 |  |  |

## 4. DISCUSSION

In this study, we reported the comparative analysis of laboratory screening for *SARS-CoV-2* among three primer-probe sets in two different master mixes (Invitrogen™ SuperScript™ III One-Step RT-PCR, and LightCycler Multiplex RNA Virus Master). The initial analysis showed the combination of TIB-Molbiol, IDT, and Phu Sa primer-probe sets were quite sensitive to positive results (4.8 copies/reaction) among Invitrogen™ SuperScript™ III One-Step RT-PCR System (Table 7). In terms of LightCycler Multiplex RNA Virus Master, TIB-Molbiol, IDT, and Phu Sa primer-probe sets also showed the same sensitivity (4.8 copies/reaction). The R-square of each primer-probe set among the different master mixes were around 0.97, the findings were compatible with a previous study [12]. Also, the values of AE in each primer-probe set among different master mixes reach the accepted criteria of AE ranged from 90-110% [11]. These findings could also contribute to gain better understandings of the combination of the best reagents for *SARS-CoV-2* screening to select the most optimum reagents to effectively halt COVID-19.

**Table 7.**
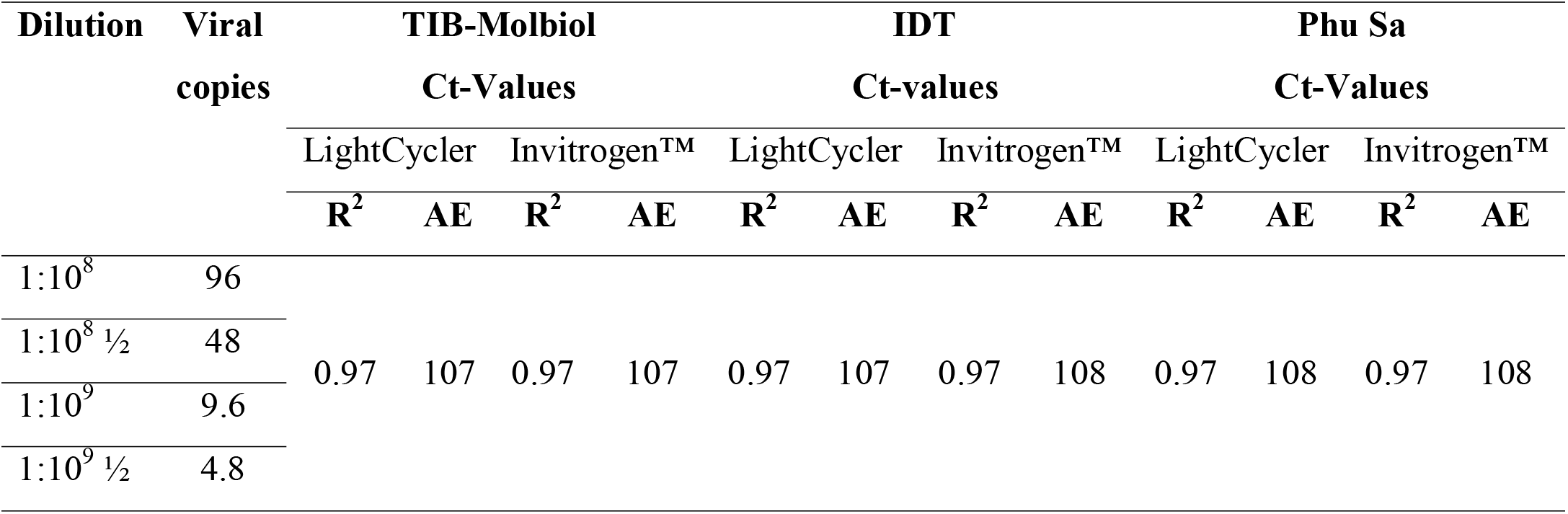
Summary of the criteria for three primer-probe sets.

The need for the optimum strategies aims to enhance testing capacity is a prerequisite because there has been an increasing figure of false positives results of RT-qPCR that were reported in the performs of *SARS-CoV-2* diagnosis for recovering patients and asymptomatic infected patients [3]. Several recent studies reported the analytical sensitivity and efficiency comparisons of *SARS-COV-2* detection by several molecular assays with multiple primer-probe sets [3, 12-14]. Although RT-qPCR is appropriate for the large-scale diagnosis of viral infection in normal viral load specimens, the RT-qPCR performance of each primer-probe set is different from others, and various primer-probe sets have contextual amplification with *SARS-CoV-2* negative nasopharyngeal swabs [4], resulting in the inconclusive results. Because of the different performances of each primer-probe set in RT-qPCR, the optimization for the design or selection of the proper primer-probe set, the appropriate annealing temperature, the limit of detection for the negative threshold determination could help to eliminate false positives or negatives once designed for clinical diagnosis of viral infection [3].

On the other hand, RT-qPCR assays targeting *SARS-CoV-2* depended on the high similarity of *SARS-CoV-2* to *SARS-CoV* as cross-react occurred. Furthermore, the less sensitivity of the assay when collecting specimens in the early time points after admission[12]. Consequently, almost all laboratories should locally validate diagnostic sensitivities and limit of detection values when beginning these assays [13].

A recent study showed that the droplet digital PCR method could diminish the inaccurate results in the low viral load specimens compared to RT-qPCR[15]. However, the possible false-positive results or false-negative results arising from current standard RT-qPCR detection of *SARS-CoV-2*, therefore, the improved choice of clinical practice could be a comprehensive approach about molecular diagnosis, X-ray or computed tomography scan, serology test, along with the decision by experienced clinicians in place of exclusively depending on RT-qPCR[15].

Several previous studies have also shown that the selection of the best primer-probe sets and equipment for *SARS-CoV-2* screening and diagnosis was an urgent and important solution for prevention and control COVID-19 [12, 14]. Our findings found that these primer-probe sets in two different master mixes were sensitive and reliable for laboratory screening of *SARS-CoV-2*. Hence, these primer-probe sets could be beneficial for the laboratory screening of *SARS-CoV-2* by RT-qPCR assay of E gene. It is crucial to improve the capacity of suspected case screenings and to reduce the affected performance of the testing that yield false-negative results [16]. The results of this study are a prelude for other studies to improve the testing capacity of screening suspected cases.

## 5. CONCLUSION

Our findings indicate TIB-Molbiol, IDT, and Phu Sa primer-probe sets could be beneficial for the laboratory screening of *SARS-CoV-2* by RT-qPCR assay of E gene. There is a need for considering the combination of these reagent sets as a new strategy to increase the testing capacity for screening programs for COVID-19.

## Data Availability

The data used to support the findings of this study are available from the corresponding author
upon request.

## Data Availability

The data used to support the findings of this study are available from the corresponding author upon request.

## Conflict interest

The authors have no conflicts of interest.

## Acknowledgments

We would like to thank colleagues at Pasteur Institute, Ho Chi Minh City for supporting the study. Thanks to Mr. Alan Tan, Sydney, Australia edited and proofread the manuscript.

## Declaration of Funding

This research received no external funding.

Abbreviation
RT-qPCR: Quantitative reverse transcription PCR
*SARS-CoV-2:*: Severe Acute Respiratory Syndrome Coronavirus 2
COVID-19: Corona virus-infected diseases 19

**Supplementary table.**
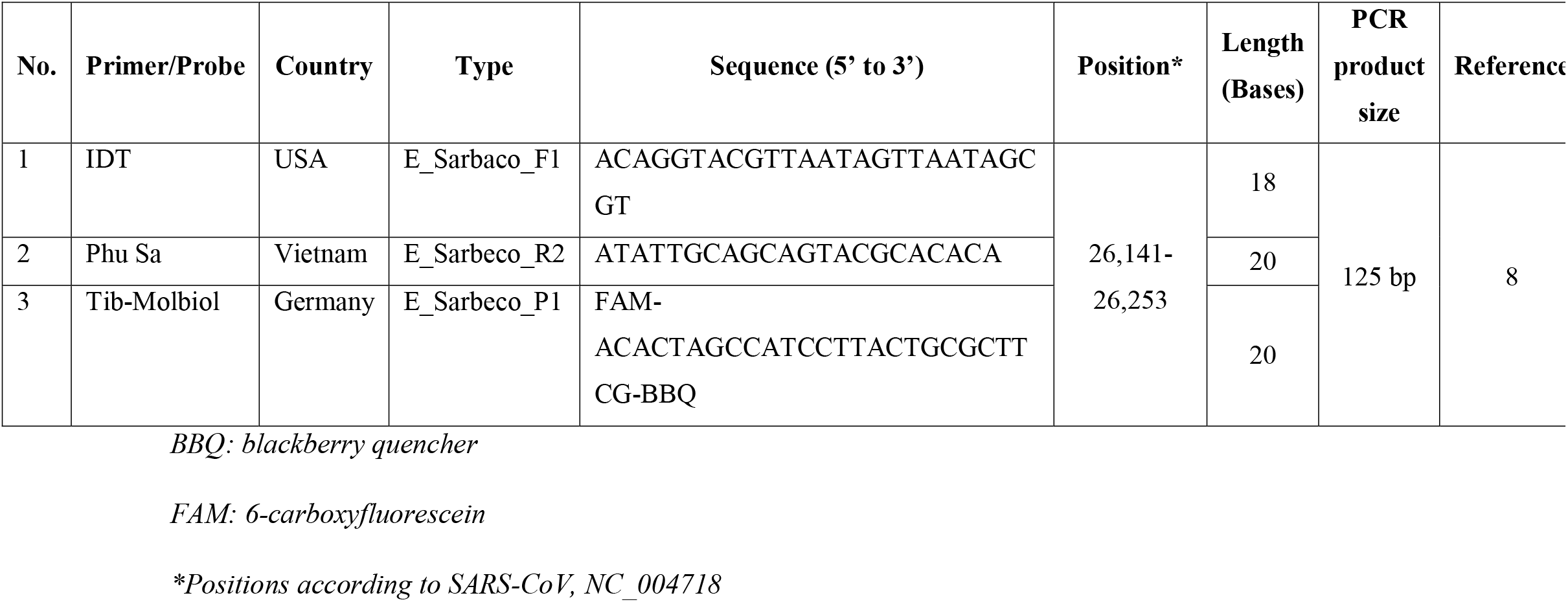
Information on primers and probes were used in this study.

## Notes

### Competing Interest Statement

The authors have declared no competing interest.

### Author Declarations

Study protocol has been reviewed and ratified by the Pasteur Institute Ho Chi Minh City Institutional Review Board (reference number: 433/XN-PAS). The clinical trial involved have been registered.

